# Homeschooling Trends Before and After the New York State Repeal of Nonmedical Vaccination Exemptions

**DOI:** 10.1101/2025.08.11.25333447

**Authors:** John W. Correira, Kathryn T. Morrison, Margaret K. Doll

## Abstract

**Importance:** Evaluation of New York State (NYS) Senate Bill 2994A repealing school-entry nonmedical vaccine exemption options suggests that the law was effective to increase school vaccine coverage; however, the law’s impact on homeschooling has not been examined.

**Objective:** To evaluate the impact of NYS Senate Bill 2994A on homeschooling prevalence.

**Design, Setting, and Participants:** In this population-based cohort study with interrupted time-series analyses, we estimated changes in homeschooling prevalence following NYS Senate Bill 2994A implementation. The study cohort comprised school districts that submitted annual student enrollment and homeschooling reports for all school years during the study period, 2014-15 through 2019-20. Analyses were conducted in January 2025.

**Exposure:** NYS Senate Bill 2994A went into effect in June 2019. Because legislative compliance was not evaluated for most students until the following school year, we considered the 2019-20 school year as the law’s effective date.

**Main Outcomes and Measures:** We calculated homeschooling prevalence as the number of homeschooling students divided by the total number of students, multiplied by 100. We estimated homeschooling prevalence differences (PD) comparing the time periods before (referent) and after NYS Senate Bill 2994A. Crude and adjusted PDs accounting for longitudinal homeschooling trends were estimated at the population-level, district-level, and county-level. Due to data limitations, PDs for New York City (NYC) were estimated only at the population-level.

**Results:** Among 685 (99.3%) NYS school districts, the repeal of nonmedical vaccine exemptions was associated with an overall increase in homeschooling prevalence of 0.1% (95% CI: 0.1%-0.1%) among NYC students and 0.3% (95% CI: 0.2%-0.3%) among students outside of NYC, after adjustment for longitudinal trends. At the district-level, the law was associated with an average 0.4% (95% CI: 0.3%-0.5%) increase in homeschooling prevalence among non-NYC schools. Spatial variation in crude homeschooling PDs was observed in county-level estimates (range: -0.3% to 1.5%; interquartile range: 0.2% to 0.5%).

**Conclusion and Relevance:** We found evidence that NYS Senate Bill 2994A was associated with small, but significant increases in homeschooling prevalence. These results suggest that a small number of un(der)vaccinated students may have disenrolled from traditional “brick-and-mortar” schools to avoid compliance with the law.

## Introduction

School entry vaccine mandates play a critical role in the assurance of high pediatric vaccine coverage in the United States (US).^1,2^ While all US states have adopted school entry vaccine mandates, most permit nonmedical exemptions for religious or personal beliefs. However, in recent years, increases in the uptake of student nonmedical vaccine exemptions have prompted some states to reconsider these policies.^3^ To address increasing nonmedical exemptions and two large measles outbreaks, New York State (NYS) passed Senate Bill 2994A repealing school entry nonmedical vaccine exemption options at public and nonpublic schools in June 2019.^4–6^

To date, evaluations of NYS Senate Bill 2994A and similar legislation in other states suggest that a repeal of nonmedical vaccine exemptions can be effective to increase school vaccination coverage;^7,8^ however, these analyses only account for students educated at “brick and mortar” schools. Where parents of un(der)vaccinated students sought alternative educational settings, such as homeschooling, to avoid immunization compliance, the effectiveness of these laws to curb vaccine-preventable disease transmission in the broader population may be compromised. Indeed, previous research has shown that vaccine mandates have been cited by some parents as salient factors in decisions to homeschool their children.^9^ Furthermore, in a survey of NYS schools, some administrators reported school enrollment declines following the adoption of NYS Senate Bill 2994A, which may represent disenrollment of un(der)vaccinated students as a mechanism to avoid compliance.^10^ Yet, in California, a legislative repeal of nonmedical vaccine exemptions was not associated with changes in homeschooling populations.^11^

To our knowledge, the impact of the NYS repeal of nonmedical vaccine exemptions on homeschooling has not yet been evaluated. Since disenrollment of un(der)vaccinated students from traditional “brick-and-mortar” schools may impact vaccine-preventable disease transmission in the broader population, we examined the relationship between Senate Bill 2994A and homeschooling prevalence in a population-based cohort of NYS school districts.

## Methods

### Study Setting

On June 13, 2019, NYS Senate Bill 2994A was signed into law repealing school-entry nonmedical vaccine exemption options from public and nonpublic schools. Students were required to demonstrate compliance within 14 days of the law’s adoption to attend school; however, due to the NYS school summer recess, compliance for most students was not assessed until the start of the 2019-20 school year.^6^

In NYS, parents who wish to homeschool children 6 to 16 years of age must file an annual notice of intent with the public school district of their residence; this notice is due either by the start of the school year (July 1), or within 14-days of the initiation of the homeschooling period.^12^ School districts are required to report homeschooling and school enrollment data to the New York State Education Department (NYSED) annually, with student counts based on the first Wednesday in October each year.^13^

### Study Design

To examine the relationship between Senate Bill 2994A and the annual prevalence of NYS homeschooling students, we conducted interrupted time-series analyses using publicly available NYSED data from the 2014-15 through 2019-20 school years. This study period was selected in consideration of the first year of data availability (2014-15) and the onset of the COVID-19 pandemic, which interrupted in-person instruction for most NYS schools beginning in March 2020. For study analyses, we considered the 2019-20 school year as the effective date for NYS Senate Bill 2994A. While school years after 2019-20 were not evaluated in relation to the repeal, we present data from the 2020-21 through 2022-23 school years, or the last year of publicly available data, for additional context regarding NYS homeschooling trends.

### Data Sources & Study Population

These analyses used publicly available, district-level homeschooling and school enrollment data files published annually on the NYSED Information and Reporting Services website.^13^ Briefly, NYSED releases separate files containing counts of public school, nonpublic school, and homeschooling students aggregated by the student’s public school district of residence. For homeschooling and nonpublic school students, these data are further stratified by age category (*i.e.,* kindergarten [K] through 6^th^ and 7^th^ through 12^th^ grades), whereas public school data are reported by grade.^13^ Using unique NYS public school district identifiers, we merged public school, nonpublic school, and homeschooling data to create a population denominator of students (*i.e.,* public, nonpublic, and homeschooling students) for each public school district area. Districts were eligible for study inclusion if annual NYSED homeschooling and school enrollment data were available for all school years in the 6-year study period (*i.e.,* 2014-15 through 2019-20).

To examine spatial homeschooling trends, we utilized publicly available county boundary data from the NYS Geographic Information System (GIS) Clearinghouse.^14^ School districts were assigned to counties using the district’s county identified in NYSED datasets.

### Statistical Analyses

We examined annual homeschooling prevalence as our outcome of interest. In separate analyses, this outcome was estimated at the: (i) population-level, representing the statewide prevalence of homeschooling; (ii) district-level, representing homeschooling prevalence by district; and (iii) county-level, representing homeschooling prevalence by county. Within each level, homeschooling prevalence was calculated by dividing the number of homeschooling students by the total number of students, multiplied by 100. For New York City (NYC), only population-level data were examined since nonpublic school enrollment was not reported by district of student residence for all study years.

Annual homeschooling prevalence was examined in relation to the adoption of Senate Bill 2994A. Since we regarded absolute differences as more meaningful than relative comparisons, we estimated homeschooling prevalence differences (PD) using the time period before the law as the referent period. In population-level analyses, the PD represented the absolute population change in homeschooling prevalence following the law. In district- and county-level analyses, the PD represented an average of PDs across units, with each unit (district or county) weighted equally.

In crude analyses, we estimated PDs and 95% confidence intervals (CIs) by directly comparing the 2019-20 versus 2018-19 (referent) school years. In adjusted analyses, statistical models included a continuous, linear term for school year (*i.e.*, 1 through 6) to adjust for longitudinal homeschooling trends; PDs and 95% CIs were estimated using a binary term to represent Senate Bill 2994A, coded as 0 and 1 in the school years pre- (2014-15 to 2018-19) and post- (2019-20) implementation, respectively. At the population-level, adjusted analyses were implemented using binomial generalized linear models (GLMs) with an identity link function and weights to account for the annual number of students under surveillance at each time point. Given potential differences in homeschooling effects, we implemented stratified GLMs, first for NYC and non-NYC populations, and next, adding age category (*i.e.*, K-6^th^ and 7-12^th^ grade students). In adjusted district- and county-level analyses (excluding NYC), binomial generalized estimating equation (GEE) models with an identify link function were used to account for repeated sampling of analysis units; stratified GEE models were implemented to examine the impact of the law on homeschooling for each age category (i.e., K-6^th^ and 7-12^th^ grade students).

In all analyses, PDs with 95% CIs that did not cross 0 were regarded as statistically significant. Analyses were conducted in January 2025 using RStudio with R version 4.2.2 (R Project for Statistical Computing).

### Ethics and Reporting

Since only publicly available, aggregate data were used in analyses, this research was exempt from ethics review and informed consent requirements under the Common Rule. We followed the Strengthening the Reporting of Observational Studies in Epidemiology (STROBE) guidelines.^15^

## Results

### Study Population and Homeschooling Frequencies

We identified 690 school districts within NYSED data, with 689 districts located outside of NYC and 1 district representing aggregated NYC student enrollment. Of these, 685 (99.3%) districts (including NYC) submitted NYSED homeschooling and school enrollment reports for all study years (*i.e.,* 2014-15 through 2019-20) and were eligible for inclusion.

Among eligible districts, a mean of 2,892,689 students (95% CI: 2,852,476-2,932,902) were included in the study population annually, with 1,728,247 students (95% CI: 1,707,159-1,749,334) from districts outside of NYC and 1,164,443 (95% CI: 1,145,076-1,183,809) from NYC. On average, 53.0% (95% CI: 52.8-53.3%) of students were enrolled in K-6^th^ grades annually. Among non-NYC school districts, the mean enrollment per district was 2,527 students (95% CI: 2,271-2,783).

Figure 1 examines frequencies of homeschooling students by year and region. Briefly, across the 6-year study period, the number of statewide K-12^th^ grade homeschooling students increased from 24,317 in the 2014-15 school year to 26,792 in the 2018-19 school year prior to the repeal. In the 2019-20 school year following Senate Bill 2994A implementation, 33,002 K-12^th^ grade students were homeschooled, representing an increase of 6,210 students from the previous year.

**Figure 1.**
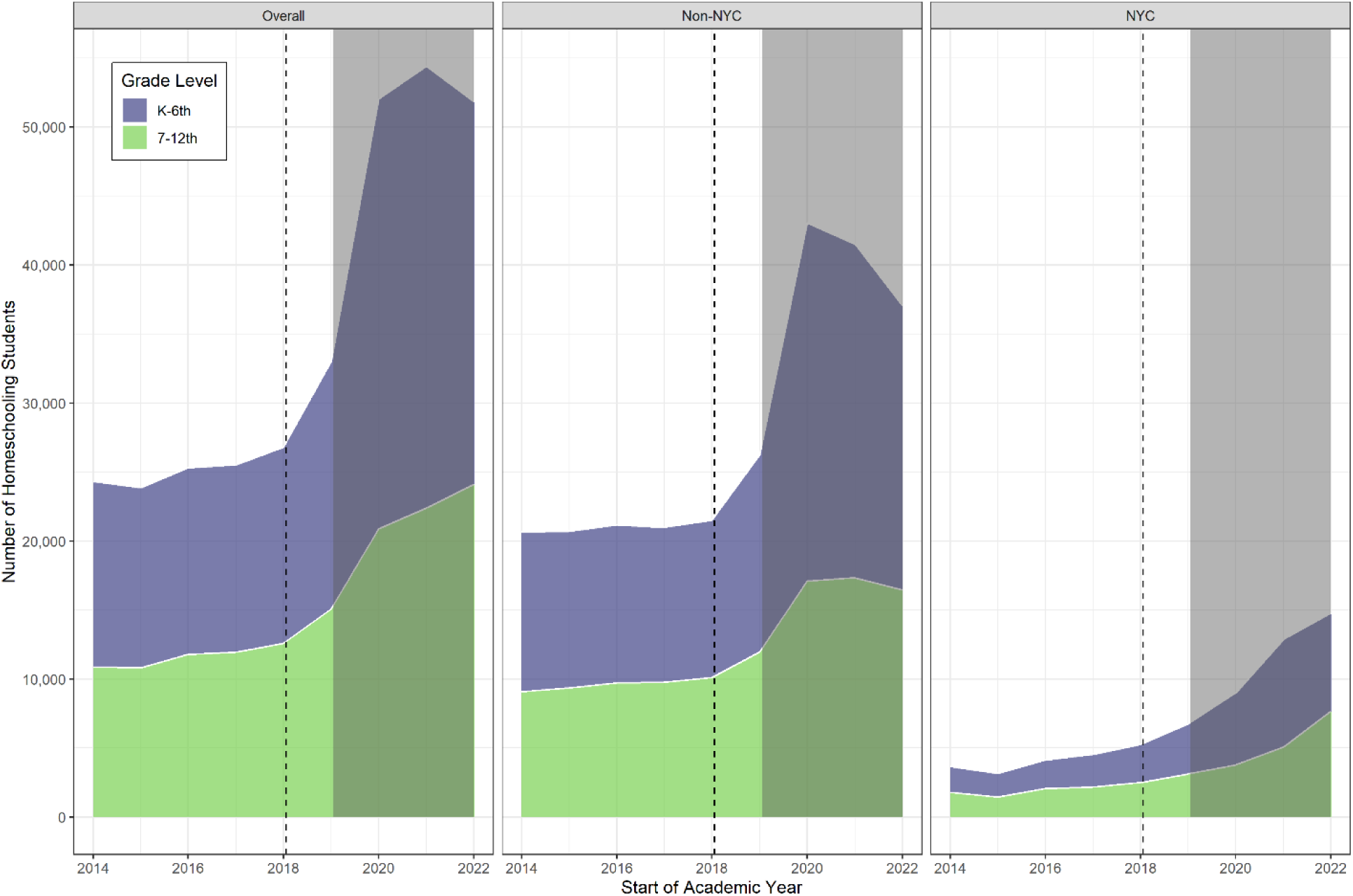
New York State homeschooling student frequencies before and after the June 2019 repeal of school-entry nonmedical vaccine exemptions. *Legend:* NYC = New York City; K-6^th^ = kindergarten through 6^th^ grade; 7-12^th^ = 7 through 12^th^ grade. *Note: The dashed line represents the school year before the implementation of New York State Senate Bill 2994A in June 2019. The shaded area represents school years affected by the COVID-19 pandemic*.

Following the onset of the COVID-19 pandemic, the number of homeschooling students increased sharply to 52,033 in the 2020-21 school year, followed by an increase to 54,399 in 2021-22; although NYS homeschooling totals in 2022-23 decreased to 51,840, the number of students remained 57% higher compared with the 2019-20 school year. Trends in homeschooling frequencies in the post-pandemic period varied by region, with the number of homeschooling students in NYC increasing across all three post-pandemic years; outside of NYC, trends were similar to the overall population.

### Population-Level Homeschooling Prevalence

Crude and adjusted population-level homeschooling prevalence estimates are displayed by region in Figure 2. In the 2018-19 school year prior to the repeal, homeschooling prevalence outside of NYC was 1.3% (95% CI: 1.2-1.3%) compared with 0.5% (95% CI: 0.4-0.5%) in NYC. Upon Senate Bill 2994A implementation, homeschooling prevalence increased to 1.5% (95% CI: 1.5-1.6%) outside of NYC and 0.6% (95% CI: 0.6-0.6%) in NYC, representing crude increases of 0.3% (95% CI: 0.3-0.3%) and 0.1% (95% CI: 0.1-0.2%), respectively, compared with the previous school year. After accounting for longitudinal homeschooling trends, results from adjusted population-level analyses were similar to crude analyses (Figure 2; eFigure 1). In adjusted analyses, Senate Bill 2994A was associated with increases of 0.3% (95% CI: 0.2-0.3%) and 0.1% (95% CI: 0.1-0.1%) in homeschooling prevalence across school districts located outside and within NYC, respectively.

**Figure 2.**
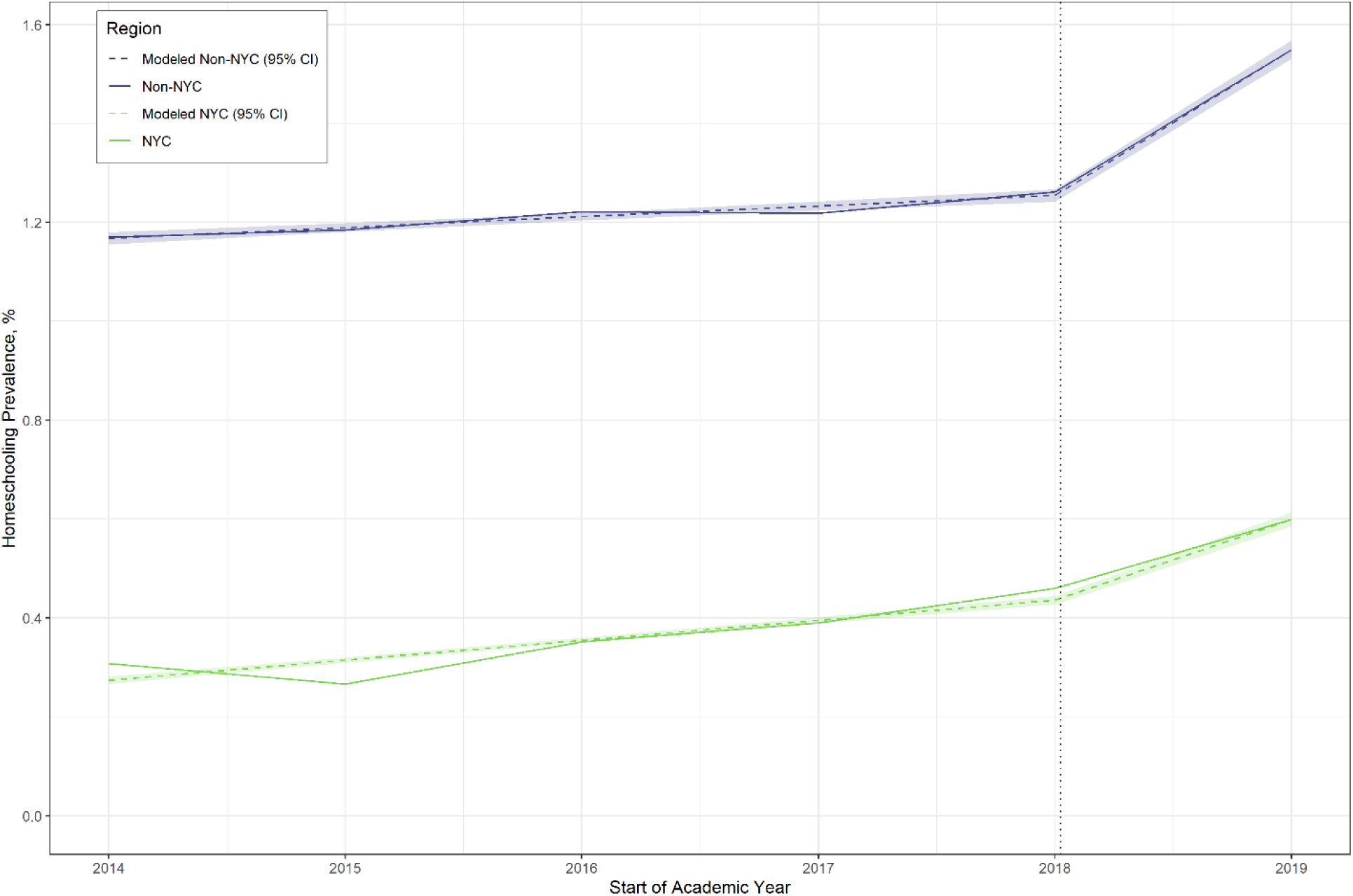
Annual crude and generalized linear modeled (GLM) estimates of population-level homeschooling prevalence in relation to the implementation of Senate Bill 2994A. *Legend*: NYC = New York City; CI = confidence interval. *Note: The dashed line represents the school year before the implementation of New York State Senate Bill 2994A in June 2019.*

Figure 3 and eFigure 2 display crude and adjusted population-level homeschooling prevalence estimates stratified by age category (i.e., K-6^th^ and 7-12^th^ grade students) and region. Upon implementation of Senate Bill 2994A, adjusted homeschooling prevalence outside of NYC increased by 0.3% (95% CI: 0.3-0.4%) among K-6^th^ grade students and 0.2% (95% CI: 0.2-0.2%) among 7-12^th^ grade students. Among K-6^th^ and 7-12^th^ grade students in NYC, adjusted homeschooling prevalence increased by 0.1% (95% CI: 0.1-0.2%) and 0.1% (95% CI: 0.1-0.1%), respectively.

**Figure 3.**
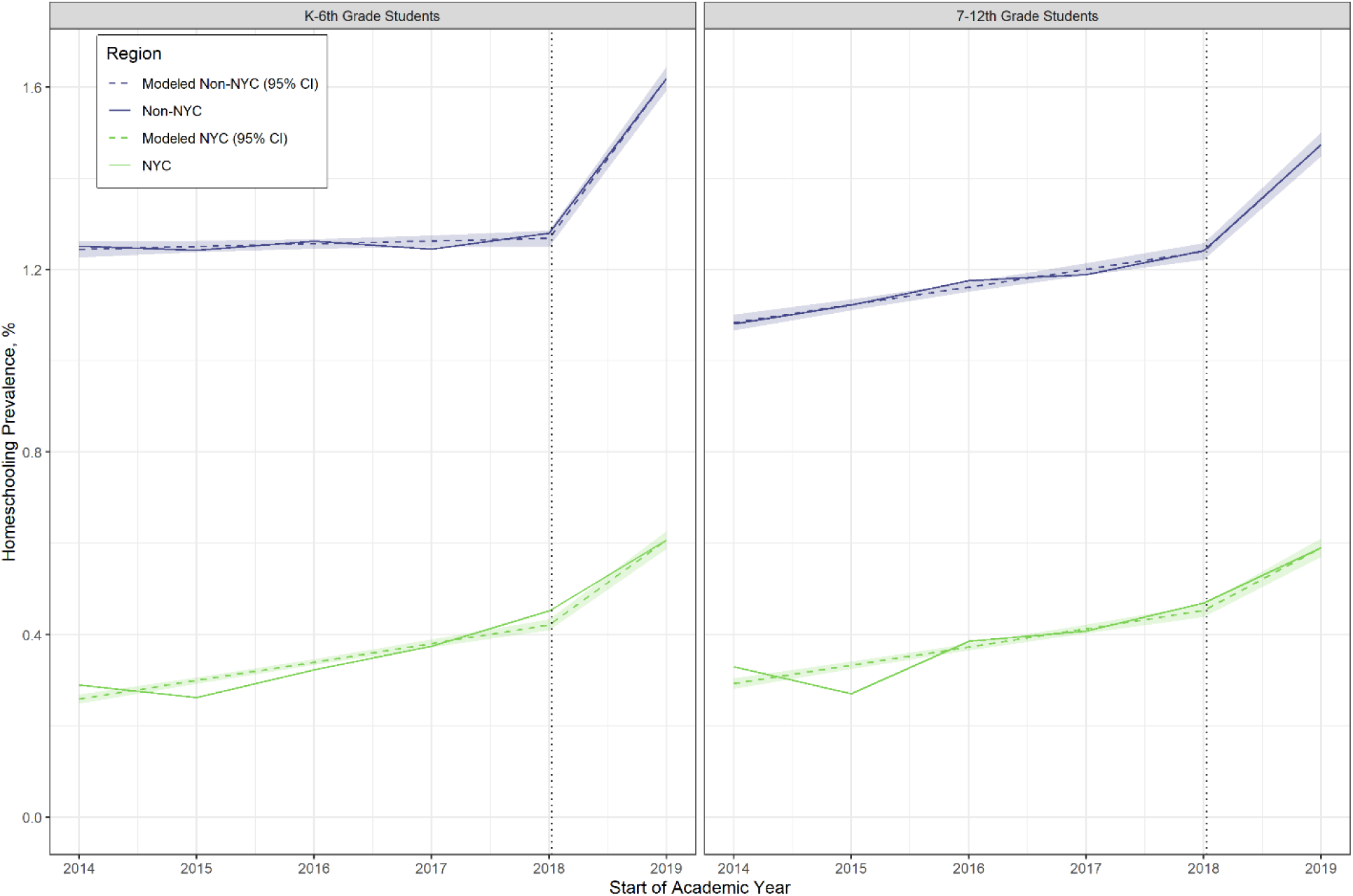
Annual crude and generalized linear modeled (GLM) population-level estimates of homeschooling prevalence stratified by student age category in relation to the implementation of Senate Bill 2994A. *Legend:* NYC = New York City; CI = confidence interval; K-6^th^ = kindergarten through 6th grade; 7-12^th^ = 7^th^ through 12^th^ grade. *Note: The dashed line represents the school year before the implementation of New York State Senate Bill 2994A in June 2019.*

### District-Level Homeschooling Trends

Figure 4 displays crude and adjusted district-level estimates of NYS homeschooling prevalence across the study period. In the year prior to Senate Bill 2994A implementation, mean homeschooling prevalence per district was 2.1% (95% CI: 1.9-2.2%) and increased to 2.5% (95% CI: 2.3-2.6%) in the 2019-20 school year, representing a mean crude PD of 0.4% (95% CI: 0.2-0.6%). Adjusted district-level estimates were similar to crude PD estimates (PD: 0.4%, 95% CI: 0.3-0.5%; eFigure 3). When stratified by student age category, the adjusted district-level mean PD was 0.5% (95% CI: 0.3-0.6%) among K-6^th^ grade students while no changes in homeschooling were observed among 7-12^th^ grade students (PD: 0.05% (95% CI: -0.3-0.4%) (eFigure 4).

**Figure 4.**
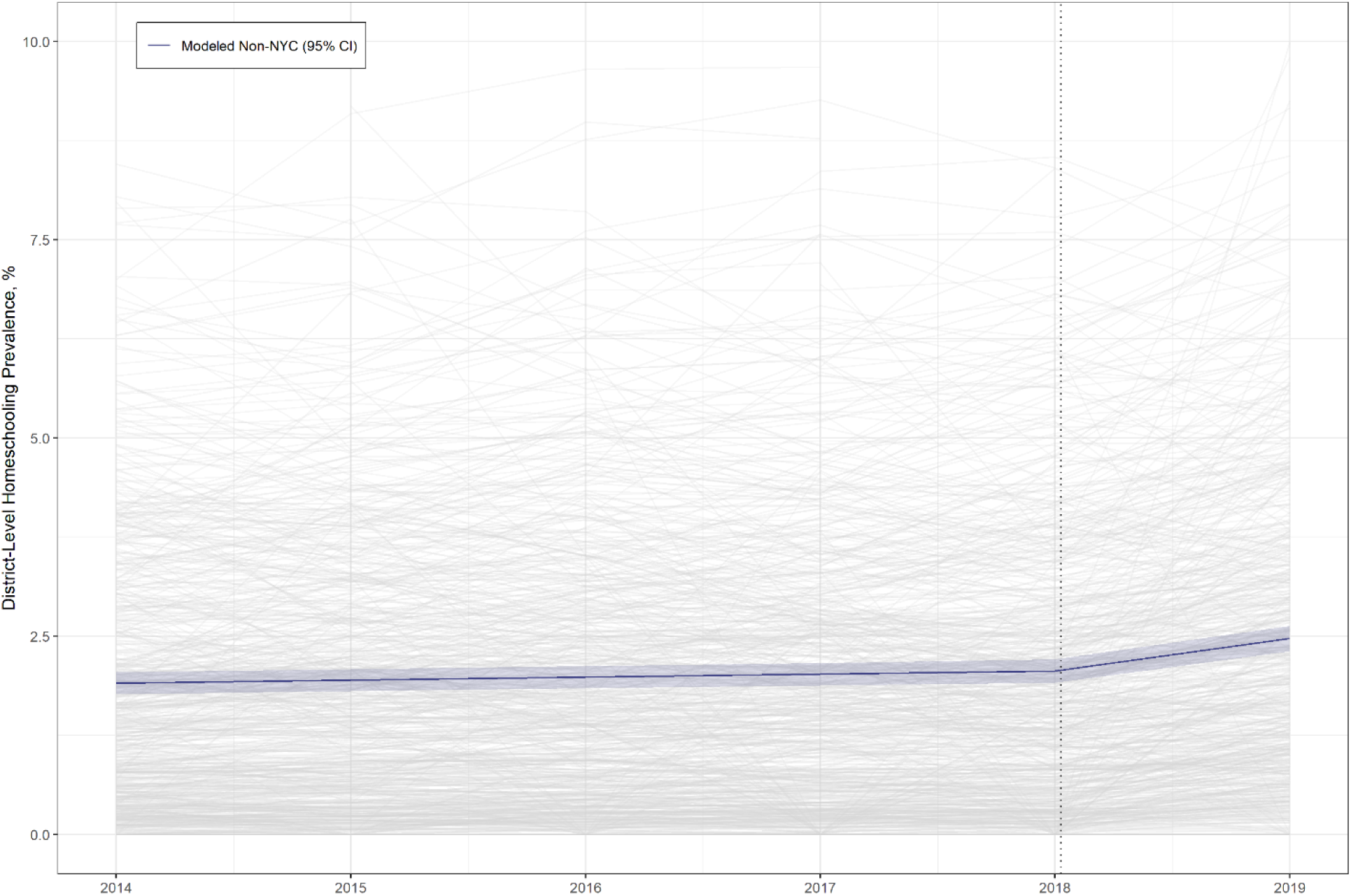
Annual crude and generalized estimating equation (GEE) modeled district-level estimates of homeschooling prevalence in relation to the implementation of Senate Bill 2994A. *Legend*: NYC = New York City; CI = confidence interval. *Note: The dashed line represents the school year before the implementation of New York State Senate Bill 2994A in June 2019. Districts with homeschooling prevalence above 10% are not displayed*

### County-Level Homeschooling Prevalence

Figure 5 displays a map of crude county-level PDs following the implementation of Senate Bill 2994A. In the year prior to Senate Bill 2994A, mean county-level homeschooling prevalence was 2.5% (95% CI: 2.1-2.8%) and increased to 2.8% (95% CI: 2.5-3.2%) in the 2019-20 school year; crude county PDs ranged from -0.3% to 1.5% (interquartile range [IQR] = 0.2 to 0.5%). The adjusted county-level mean PD associated with the law was similar to crude PD estimates, with an increase of 0.3% (95% CI: 0.3-0.4%; eFigure 5) observed following implementation. In adjusted analyses stratified by student age category, increases of 0.4% (95% CI: 0.3-0.5%) among K-6^th^ grade and 0.3% (95% CI: 0.1-0.4%) among 7-12^th^ grade students were observed (eFigure 6). Longitudinal homeschooling prevalence data by county are displayed in eFigure 7.

**Figure 5.**
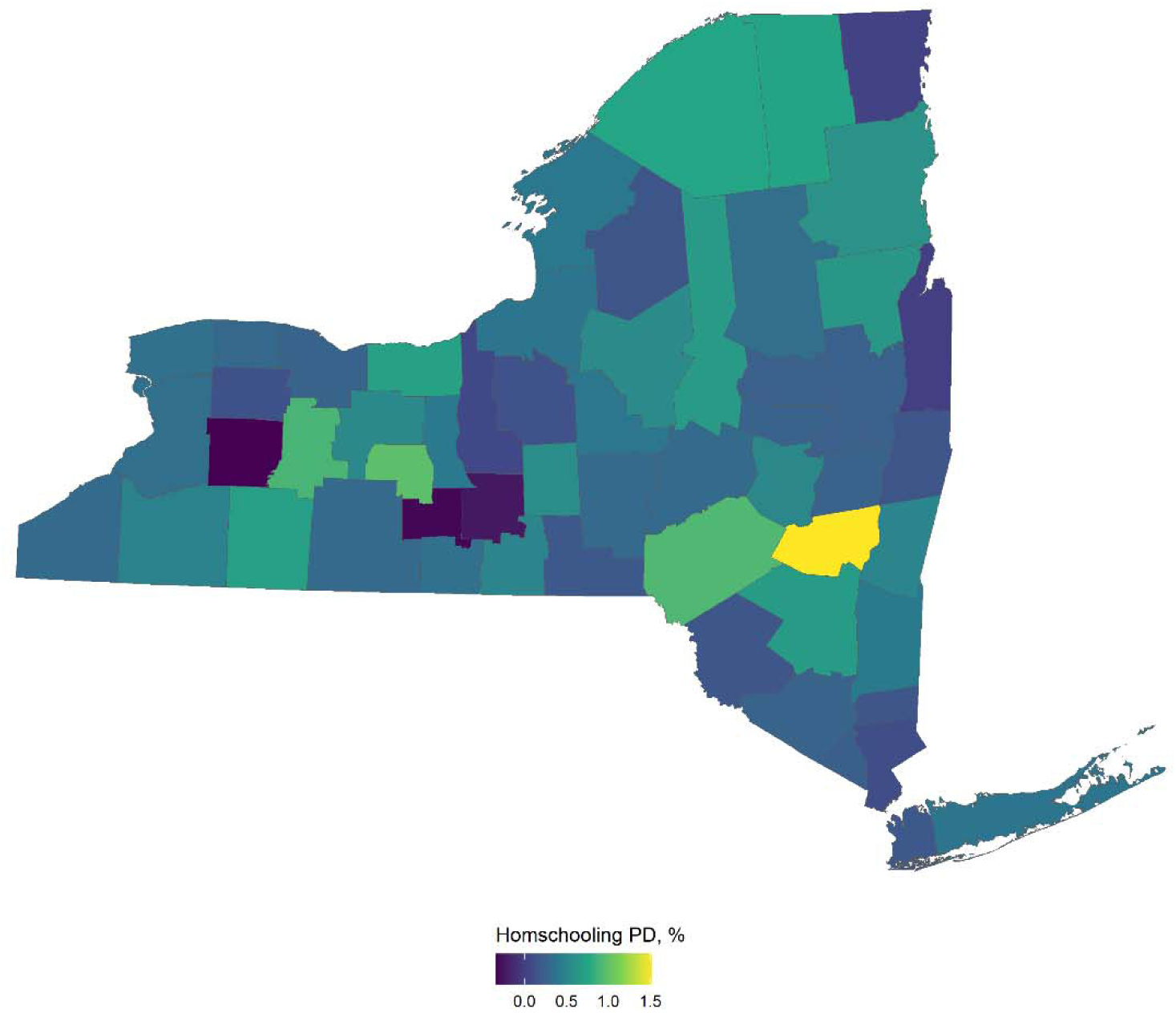
County-level crude homeschooling prevalence differences (PD) comparing the year after versus before (referent) the New York State repeal of school-entry nonmedical vaccination exemptions. *Legend*: PD = prevalence difference.

## Discussion

In this population-based cohort study with interrupted time-series analyses, we evaluated the impact of the NYS repeal of school-entry nonmedical vaccine exemptions on homeschooling prevalence. Following the adoption of Senate Bill 2994A, we observed small, but significant increases in population-level homeschooling prevalence, with increases of 1 homeschooling student per 1,000 students (0.1%) in NYC and 3 homeschooling students per 1,000 students (0.3%) outside of NYC. In both regions, increases were larger among K-6^th^ grade (NYC: 0.1%; non-NYC: 0.3%) versus 7-12^th^ grade students (NYC: 0.1%; non-NYC: 0.2%). In districts outside of NYC, we also estimated that the law was associated with a mean increase of 4 homeschooling students per 1,000 district students (0.4%); however, when stratified by student age category, increases in homeschooling were observed only among K-6^th^ grade students (0.5%), while trends among 7-12^th^ grade students remained stable.

Because our analyses accounted for longitudinal homeschooling trends, we hypothesize that the changes we observed likely reflect homeschooling of un(der)vaccinated students to avoid legislative compliance. Should this hypothesis be correct, increases in homeschooling prevalence represent a small offset to broader NYS pediatric vaccine coverage gains attributable to the law. Nonetheless, evaluation of the law’s effectiveness in a population-based study of non-NYC schools found that the repeal was successful to increase school vaccine coverage by nearly 6% and 1% at nonpublic and public schools, respectively.^7^ Together with our results, these data suggest the law was likely effective to drive meaningful, net positive vaccine coverage gains in the broader NYS pediatric population as well.

New York is one of several states that recently repealed nonmedical vaccine exemption options from school-entry mandates.^6,16–18^ In California, a similar legislative repeal was not associated with changes in the homeschooling population;^11^ however, unlike in NYS,^7^ increases in student medical exemptions were observed.^8^ We hypothesize that these differing experiences may be due to state-level variation in the designs and implementation timelines of the legislative repeals. Indeed, NYS Senate Bill 2994A became effective immediately and did not include a grandfather clause excusing students with existing nonmedical vaccine exemptions from compliance;^6^ in contrast, California’s repeal included a grace period between the law’s passage and its implementation, while also allowing students with existing nonmedical exemptions to retain such exemptions until reaching a grade requiring compliance re-evaluation (*i.e.,* K or 7^th^ grades).^18^ These differences may have placed greater pressure on parents of un(der)vaccinated NYS students following the law’s passage, making them more likely to homeschool their children as a mechanism to avoid compliance. Policymakers may wish to consider these contrasting outcomes and their public health implications, such as susceptibility to vaccine-preventable diseases and outbreaks,^19,20^ when weighing the adoption of similar legislation.

Like California,^8,21^ our results suggest potential spatial variation in the impact of NYS Senate Bill 2994A. Specifically, county-level changes in homeschooling prevalence ranged from -0.3% to 1.5% following the repeal of nonmedical vaccine exemptions. Despite high “brick-and-mortar” school-level vaccine coverage, this spatial heterogeneity suggests that some NYS communities may be differentially susceptible to vaccine-preventable disease outbreaks due to clustering of un(der)vaccinated residents.^19,20^ Further research is needed to better understand geographic differences in the law’s effects.

Interestingly, the increases in homeschooling populations we observed in relation to NYS Senate Bill 2994A’s adoption were small compared with increases following the onset of the COVID-19 pandemic. Although data from the most recent school year examined (2022-23) suggest that NYS homeschooling populations may now be decreasing, the number of homeschooling students remained more than 50% higher than in the 2019-20 school year, or the year NYS Senate Bill 2994A went into effect. While these changes are unlikely to be related to the repeal, increases in NYS homeschooling students may present challenges for public health officials since homeschooling populations are not required to report their vaccination status nor comply with NYS school vaccination requirements. These findings underscore the importance of continued efforts by public health officials to engage homeschooling communities and to maintain trust and support for pediatric vaccinations to assure high vaccine uptake.

### Limitations

This study is subject to several limitations. First, since parents of NYS homeschooling students are not required to report their child’s vaccination status, we cannot confirm whether the changes we observed in homeschooling prevalence represent increases in un(der)vaccinated homeschooling students. Second, due to the COVID-19 pandemic, our study period was necessarily limited to one year following the implementation of Senate Bill 2994A. These limitations are partially offset by our interrupted time-series study design, which accounted for homeschooling trends in the five years before the law’s implementation when estimating changes in homeschooling prevalence. Third, the effective date of Senate Bill 2994A implementation coincided with the 2019-20 school year, which was subsequently disrupted by the COVID-19 pandemic; however, based on the timing of most NYS pandemic school restrictions (March 2020) and NYSED annual district reporting (October 2019),^13^ it is unlikely that the changes in homeschooling we observed in the 2019-20 school year were related to the COVID-19 pandemic. Fourth, since NYSED data are not available for each grade, we were unable to assess grade-level trends in homeschooling; nonetheless, we analyzed differences in homeschooling by student age category (*i.e.,* K-6^th^ vs. 7-12^th^ grade) to provide insight into differences in the law’s impact by age. Fifth, our analyses relied on NYSED administrative datasets, which were not created for this research; nonetheless, we observed high district completion of NYSED reports across the study period, enabling us to include a nearly complete (99.3%) population-based sample of NYS school districts in our analyses. Sixth, we assumed that homeschooling trends prior to the implementation of the Senate Bill 2994A were linear, which we deemed appropriate based on comparisons of crude vs. modelled data. Finally, we also assumed that changes occurring concurrently with the law’s implementation would not impact our outcome, a key assumption of the interrupted time-series study design.

### Conclusion

We found that NYS Senate Bill 2994A repealing school-entry nonmedical vaccine exemption options was associated with small, but significant increases in homeschooling prevalence. These changes suggest that a small number of un(der)vaccinated students may have disenrolled from traditional “brick-and-mortar” schools to avoid legislative compliance, likely representing a small offset to broader NYS pediatric vaccine coverage gains attributable to the law. Nonetheless, while our findings underscore the importance of examining the potential unintended effects of vaccine legislation, Senate Bill 2994A was an effective policy to increase student vaccination coverage at NYS schools.^7^

## Supporting information

Supplementatry Online Content

## Access to Data and Data Analysis

JWC and MKD had full access to all data in the study and take responsibility for the integrity of the data and the accuracy of the data analysis. KM reviewed and provided support for statistical analyses.

## Data Sharing Statement

The data used in these analyses are publicly available from the New York State Department of Health and the New York State Education Department.

