## Supplementatry Online Content for "Homeschooling Trends Before and After the New York State Repeal of Nonmedical Vaccination Exemptions"

**eFigure 1.** Population-level binomial generalized linear models (GLMs) and results estimating the relationship between New York State Senate Bill 2994A and homeschooling prevalence; stratified models were implemented by NYS region (i.e., New York City (NYC) and non-NYC).

**eFigure 2.** Population-level binomial generalized linear models (GLMs) and results estimating the relationship between New York State Senate Bill 2994A and homeschooling prevalence; stratified models were implemented by NYS region (i.e., New York City (NYC) and non-NYC) and student age category (i.e., K-6^th^ and 7-12^th^ grade).

**eFigure 3.** District-level binomial generalized estimating equation (GEE) model and results estimating the relationship between New York State Senate Bill 2994A and district-level homeschooling prevalence.

**eFigure 4.** District-level binomial generalized estimating equation (GEE) model and results estimating the relationship between New York State Senate Bill 2994A and district-level homeschooling prevalence; stratified models were implemented by student age category (i.e., K-6^th^ and 7-12^th^ grade).

**eFigure 5.** County-level binomial generalized estimating equation (GEE) model and results estimating the relationship between New York State Senate Bill 2994A and county-level homeschooling prevalence.

**eFigure 6.** County-level binomial generalized estimating equation (GEE) model and results estimating the relationship between New York State Senate Bill 2994A and county-level homeschooling prevalence; stratified models were implemented by student age category (i.e., K-6^th^ and 7-12^th^ grade).

**eFigure 7.** Annual crude and generalized estimating equation (GEE) modeled county-level estimates of homeschooling prevalence in relation to the implementation of Senate Bill 2994A.

**eFigure 1.** Population-level binomial generalized linear models (GLMs) and results estimating the relationship between New York State Senate Bill 2994A and homeschooling prevalence; stratified models were implemented by NYS region (*i.e.*, New York City (NYC) and non-NYC).

| **Binomial GLM with an Identity Link Function:**  ${Homeschool}_{j}/{Total}_{j}=\beta_{0}+\beta_{1}{Time}_{j}+\beta_{2}{Law}_{j}+\varepsilon_{j}$ | | | | |
| --- | --- | --- | --- | --- |
| **Term** | **Estimate** | **P value** | **Lower CI** | **Upper CI** |
| *Stratified Model: NYC* | | | | |
| Intercept | 0.0027 | < 0.001 | 0.0027 | 0.0028 |
| Time | 0.0004 | < 0.001 | 0.0004 | 0.0004 |
| Law | 0.0012 | < 0.001 | 0.0010 | 0.0014 |
| *Stratified Model: Non-NYC* | | | | |
| Intercept | 0.0117 | < 0.001 | 0.0116 | 0.0118 |
| Time | 0.0002 | < 0.001 | 0.0002 | 0.0003 |
| Law | 0.0027 | < 0.001 | 0.0025 | 0.0030 |
| **Where:**   - *Homeschool* = Homeschool enrollment - *Total =* Total student enrollment - *Time* = Time from start of study (years) - *Law* = New York State Senate Bill 2994A implementation   (0 = pre- and 1 = post-implementation)   - $\varepsilon$ = Error term - *j =* Time   Note: Models were weighted by total student enrollment for each time point. | | | | |

**eFigure 2.** Population-level binomial generalized linear models (GLMs) and results estimating the relationship between New York State Senate Bill 2994A and homeschooling prevalence; stratified models were implemented by NYS region (i.e., New York City (NYC) and non-NYC) and student age category (i.e., K-6^th^ and 7-12^th^ grade).

| **Binomial GLM with an Identity Link Function:**  ${Homeschool}_{j}/{Total}_{j}=\beta_{0}+\beta_{1}{Time}_{j}+\beta_{2}{Law}_{j}+\varepsilon_{j}$ | | | | |
| --- | --- | --- | --- | --- |
| **Term** | **Estimate** | **P value** | **Lower CI** | **Upper CI** |
| *Stratified Model: NYC, K-6^th^ grade students* | | | | |
| Intercept | 0.0026 | < 0.001 | 0.0025 | 0.0027 |
| Time | 0.0004 | < 0.001 | 0.0004 | 0.0005 |
| Law | 0.0014 | < 0.001 | 0.0012 | 0.0017 |
| *Stratified Model: NYC, 7-12^th^ grade students* | | | | |
| Intercept | 0.0029 | < 0.001 | 0.0028 | 0.0030 |
| Time | 0.0004 | < 0.001 | 0.0003 | 0.0004 |
| Law | 0.0010 | < 0.001 | 0.0007 | 0.0013 |
| *Stratified Model: Non-NYC, K-6^th^ grade students* | | | | |
| Intercept | 0.0124 | < 0.001 | 0.0123 | 0.0126 |
| Time | 0.0001 | 0.1080 | -0.00001 | 0.0001 |
| Law | 0.0034 | < 0.001 | 0.0031 | 0.0038 |
| *Stratified Model: Non-NYC, 7-12^th^ grade students* | | | | |
| Intercept | 0.0108 | < 0.001 | 0.0107 | 0.0110 |
| Time | 0.0004 | < 0.001 | 0.0003 | 0.0005 |
| Law | 0.0020 | < 0.001 | 0.0016 | 0.0023 |
| **Where:**   - *Homeschool* = Homeschool enrollment - *Total =* Total student enrollment - *Time* = Time from start of study (years) - *Law* = New York State Senate Bill 2994A implementation   (0 = pre- and 1 = post-implementation)   - $\varepsilon$ = Error term - *j =* Time   Note: Models were weighted by total student enrollment for each time point. | | | | |

**eFigure 3.** District-level binomial generalized estimating equation (GEE) model and results estimating the relationship between New York State Senate Bill 2994A and district-level homeschooling prevalence.

| **Binomial GEE Model with an Identity Link Function:**  ${Homeschool}_{ij}/{Total}_{ij}=\beta_{0}+\beta_{1}{Time}_{ij}+\beta_{2}{Law}_{ij}+\varepsilon_{ij}$ | | | | |
| --- | --- | --- | --- | --- |
| **Term** | **Estimate** | **P value** | **Lower CI** | **Upper CI** |
| Intercept | 0.0191 | < 0.001 | 0.0177 | 0.0204 |
| Time | 0.0004 | < 0.001 | 0.0002 | 0.0006 |
| Law | 0.0037 | < 0.001 | 0.0028 | 0.0046 |
| **Where:**   - *Homeschool* = Homeschool enrollment - *Total =* Total enrollment - *Time* = Time from start of study (years) - *Law* = New York State Senate Bill 2994A implementation   (0 = pre- and 1 = post-implementation)   - $\varepsilon$ = Error term - *i =* Individual school district - *j =* Time | | | | |

**eFigure 4.** District-level binomial generalized estimating equation (GEE) model and results estimating the relationship between New York State Senate Bill 2994A and district-level homeschooling prevalence; stratified models were implemented by student age category (i.e., K-6^th^ and 7-12^th^ grade).

| **Binomial GEE Model with an Identity Link Function:**  ${Homeschool}_{ij}/{Total}_{ij}=\beta_{0}+\beta_{1}{Time}_{ij}+\beta_{2}{Law}_{ij}+\varepsilon_{ij}$ | | | | |
| --- | --- | --- | --- | --- |
| **Term** | **Estimate** | **P value** | **Lower CI** | **Upper CI** |
| *Stratified Model: K-6^th^ grade students* | | | | |
| Intercept | 0.0202 | < 0.001 | 0.0187 | 0.0216 |
| Time | 0.0001 | 0.2406 | -0.0001 | 0.0004 |
| Law | 0.0045 | < 0.001 | 0.0034 | 0.0056 |
| *Stratified Model: 7-12^th^ grade students* | | | | |
| Intercept | 0.0207 | < 0.001 | 0.0177 | 0.0237 |
| Time | 0.0013 | 0.0135 | 0.0003 | 0.0024 |
| Law | 0.0005 | 0.8056 | -0.0034 | 0.0044 |
| **Where:**   - *Homeschool* = Homeschool enrollment - *Total =* Total enrollment - *Time* = Time from start of study (years) - *Law* = New York State Senate Bill 2994A implementation   (0 = pre- and 1 = post-implementation)   - $\varepsilon$ = Error term - *i =* Individual school district - *j =* Time | | | | |

**eFigure 5.** County-level binomial generalized estimating equation (GEE) model and results estimating the relationship between New York State Senate Bill 2994A and county-level homeschooling prevalence.

| **Binomial GEE Model with an Identity Link Function:**  ${Homeschool}_{ij}/{Total}_{ij}=\beta_{0}+\beta_{1}{Time}_{ij}+\beta_{2}{Law}_{ij}+\varepsilon_{ij}$ | | | | |
| --- | --- | --- | --- | --- |
| **Term** | **Estimate** | **P value** | **Lower CI** | **Upper CI** |
| Intercept | 0.0229 | < 0.001 | 0.0199 | 0.0260 |
| Time | 0.0004 | < 0.001 | 0.0002 | 0.0006 |
| Law | 0.0033 | < 0.001 | 0.0026 | 0.0041 |
| **Where:**   - *Homeschool =* Homeschool enrollment - *Total =* Total enrollment - *Time*= Time from start of study (years) - *Law* = New York State Senate Bill 2994A implementation   (0 = pre- and 1 = post-implementation)   - $\varepsilon$ = Error term - *i =* Individual county - *j =* Time | | | | |

**eFigure 6.** County-level binomial generalized estimating equation (GEE) model and results estimating the relationship between New York State Senate Bill 2994A and county-level homeschooling prevalence; stratified models were implemented by student age category (i.e., K-6^th^ and 7-12^th^ grade).

| **Binomial GEE Model with an Identity Link Function:**  ${Homeschool}_{ij}/{Total}_{ij}=\beta_{0}+\beta_{1}{Time}_{ij}+\beta_{2}{Law}_{ij}+\varepsilon_{ij}$ | | | | |
| --- | --- | --- | --- | --- |
| **Term** | **Estimate** | **P value** | **Lower CI** | **Upper CI** |
| *Stratified Model: K-6^th^ grade students* | | | | |
| Intercept | 0.0243 | < 0.001 | 0.0210 | 0.0276 |
| Time | 0.0001 | 0.4476 | -0.0002 | 0.0004 |
| Law | 0.0039 | < 0.001 | 0.0027 | 0.0050 |
| *Stratified Model: 7-12^th^ grade students* | | | | |
| Intercept | 0.0214 | < 0.001 | 0.0185 | 0.0242 |
| Time | 0.0008 | < 0.001 | 0.0004 | 0.0011 |
| Law | 0.0027 | < 0.001 | 0.0013 | 0.0040 |
| **Where:**   - *Homeschool* = Homeschool enrollment - *Total =* Total enrollment - *Time* = Time from start of study (years) - *Law* = New York State Senate Bill 2994A implementation   (0 = pre- and 1 = post-implementation)   - $\varepsilon$ = Error term - *i =* Individual county - *j =* Time | | | | |

**eFigure 7.** Annual crude and generalized estimating equation (GEE) modeled county-level estimates of homeschooling prevalence in relation to the implementation of Senate Bill 2994A.

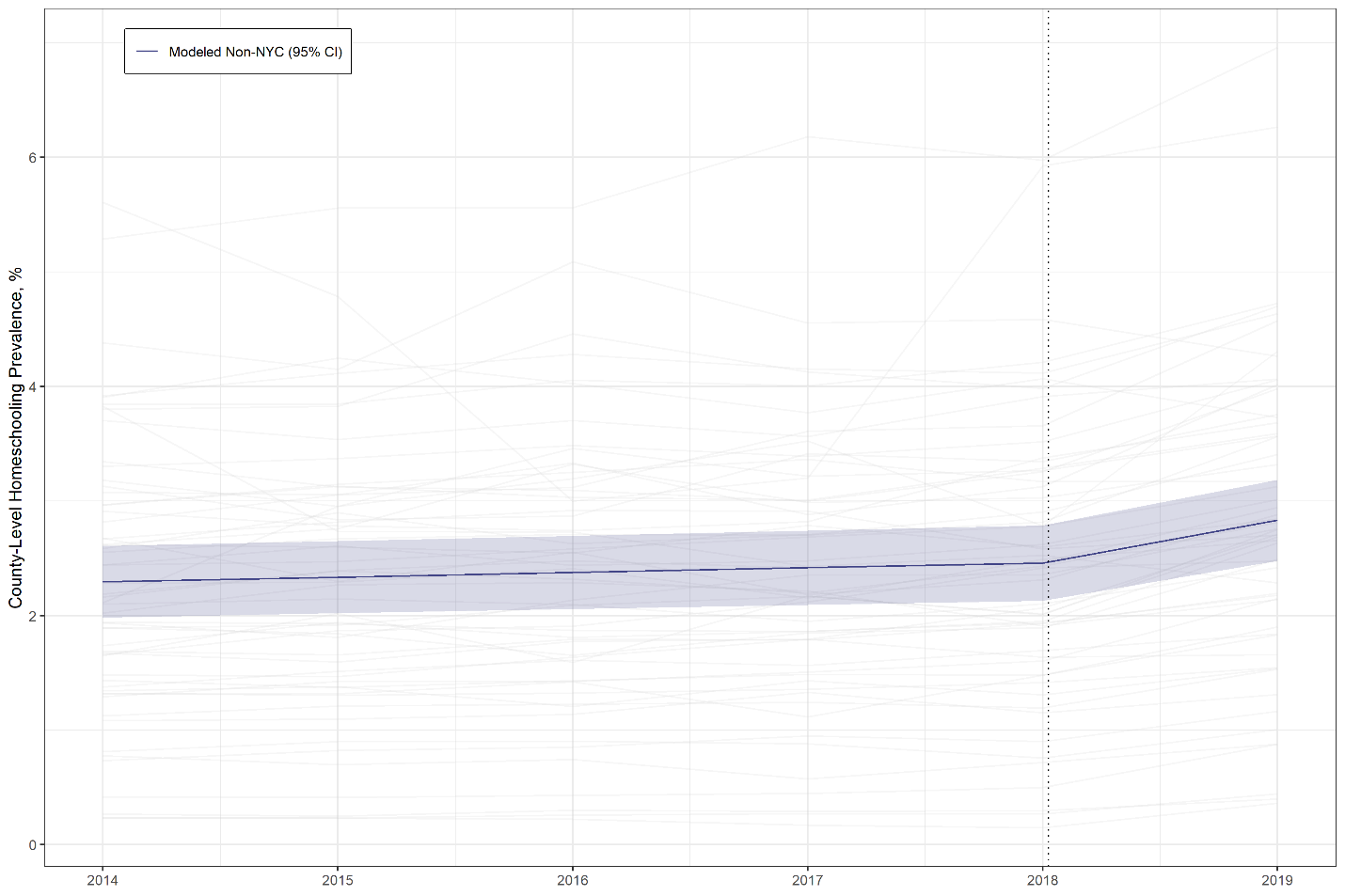

*Legend:* NYC = New York City; CI = confidence interval.

*Note: The dashed line represents the school year before the implementation of New York State Senate Bill 2994A in June 2019.*
